# Adjusting for the reduced sensitivity of CXR in the dose-response relationship between cumulative silica exposure and silicosis in miners

**DOI:** 10.1101/2025.05.29.25328501

**Authors:** Patrick Howlett, Ashwin Durairaj, Jeffery Gan, Maia Lesosky, Johanna Feary

## Abstract

**Introduction:** A recent meta-analysis confirmed that Chest Xray (CXR) has low sensitivity for the diagnosis of silicosis however the impact of this is unclear. We therefore re-estimated a previously published dose-response relationship between cumulative respirable crystalline silica (RCS) exposure and silicosis risk, under the assumptions that sensitivity was either fixed or relative to the population proportion of severe silicosis.

**Methods:** We combined unpublished logistic regression models from Scottish coal miners with meta-analysis results to model how CXR sensitivity changed according to cumulative RCS exposure. Among mining cohorts, we calculated the difference in the cumulative risk of silicosis between the unadjusted, and fixed and relative scenarios. Finally, we re-estimated the dose-response meta-analysis and related absolute risk reductions (ARR).

**Results:** In all mining cohorts the cumulative risk of silicosis was substantially higher in both the fixed and relative sensitivity scenarios compared to the unadjusted estimate. This was most pronounced in the relative scenario and when cumulative RCS exposures were below approximately 6 mg/m^3^-years. A reduction from 4 to 2 mg/m^3^-years in cumulative RCS exposure corresponded to larger ARRs in the fixed and relative scenario than the unadjusted scenario; 409 (374, 434) and 557 (451, 620) cases per 1000 miners compared to 323 (298, 344) cases per 1000 miners, respectively.

**Discussion:** We were reliant on a single estimate of the proportion of severe disease to link sensitivity and cumulative RCS exposure. Nevertheless, adjusting for the low sensitivity of CXR for silicosis results in meaningful increased risks of silicosis in previously published mining cohorts.

## Introduction

Concerns regarding a resurgence of silicosis globally have prompted reviews that attempt to determine the dose-response relationship between silica exposure and silicosis risk and disease thresholds [1–3]. All reviewed studies use Chest Xray (CXR) as the diagnostic method for silicosis. However, one of the main sources of bias has been the considerable proportion of silicosis present on high resolution computed tomography (HRCT) or autopsy that is not visible on chest Xray (CXR) [4].

We recently demonstrated the reduced sensitivity of CXR for silicosis, compared to HRCT or autopsy under two counterfactual scenarios [5]. First, assuming sensitivity of CXR was fixed, we estimated sensitivity to be 0.76 (95% confidence interval (CI) 0.63, 0.86) compared to the High-Resolution Computed Tomography (HRCT).

Second, assuming that CXR sensitivity was entirely explained by the proportion of severe silicosis in the population, our meta-regression estimated that sensitivity increased by 1.15% (95% CI 0.44, 1.86) for each 1% increase in proportion of severe silicosis (R^2^ = 63%; I^2^ = 86%). Severe silicosis was defined as the proportion of International Labour Organisation (ILO) Classification category ≥2/1 relative to ≥1/0 [5].

As silicosis risk is related to cumulative silica exposure, by extension, the sensitivity of CXR for silicosis may be related to cumulative silica exposure, either in a fixed manner or relative manner as described above. The later scenario has previously been investigated by Hnizdo et al who estimated that, for a miner with 10 years exposure, the positive predictive value of CXR increased from 14% with 1 mg/m^3^- years to 97% with 4 mg/m^3^-years [6].

Given the potential magnitude of the underestimation of silicosis cases, adjusting for the reduced sensitivity of CXR for silicosis is an important step in improving our current understanding of the dose-response relationship between cumulative silica exposure and silicosis risk [1].

In this analysis, using information from a Scottish miners cohort [7], we modelled how CXR sensitivity changes according to cumulative silica exposure. Under fixed and relative sensitivity scenarios, we then re-estimated cumulative risk and the dose-response relationship between cumulative silica exposure and silicosis among miner cohorts.

## Methods

Throughout our analysis the fixed sensitivity scenario assumes sensitivity to be 0.76 (95% CI 0.63, 0.86); the CXR sensitivity compared to the HRCT reference standard in our meta-analysis [5].

To estimate relative sensitivity, we obtained unpublished logistic regression models of silicosis risk from a study of Scottish coal miners [7,8] and calculated the proportion of CXR ILO category ≥2/1 silicosis relative to ≥1/0 according to cumulative RCS. Using these values in our previously published meta-regression, we then estimated the corresponding relative sensitivity of CXR according to cumulative RCS exposure. Example calculations are provided in Supplementary table 1.

Using point-estimates from our fixed and relative sensitivity scenarios, we re-estimated the number of silicosis cases in each category of cumulative RCS exposure of miner cohorts in our previous review [1]. We then estimated cumulative risk using the Steenland method [9]. As previously, we fitted non-parametric locally weighted least squares estimates (LOESS) to these categories and extracted the median and quartile cumulative risks at exposures of 1, 2, 4 and 10 mg/m^3^-years.

For fixed and relative sensitivity scenarios, using previous study methods [1], we re- calculated the dose-response meta-analysis and absolute risk reduction (ARR) compared to a baseline of 4 mg/m^3^-years cumulative exposure (equivalent to 40 years at 0.1 mg/m^3^). As the distribution relative risks differed across scenarios, a secondary analysis used empirical knot values of 0.5, 3.5, 8 and 15 mg/m^3^-years.

Importantly, we made two assumptions. For the cumulative risk analysis, the maximum number of cases, following adjustment for sensitivity, could not exceed the total number in the category (i.e. all had disease). For the dose-response analysis, as non-infinite risk ratios are required, the maximum possible number was the total number of cases, minus one.

Code and data are available at https://github.com/pjhowlett/da_silic_cxr/tree/main.

Human Research Ethics Committee approval was not required as previously published data was used.

## Results

Under the relative sensitivity scenario, CXR sensitivity increased from 0.29 when cumulative silica exposure was zero to 1 at 10.14 mg/m^3^-years (Supplementary figure 1).

**Figure 1.**
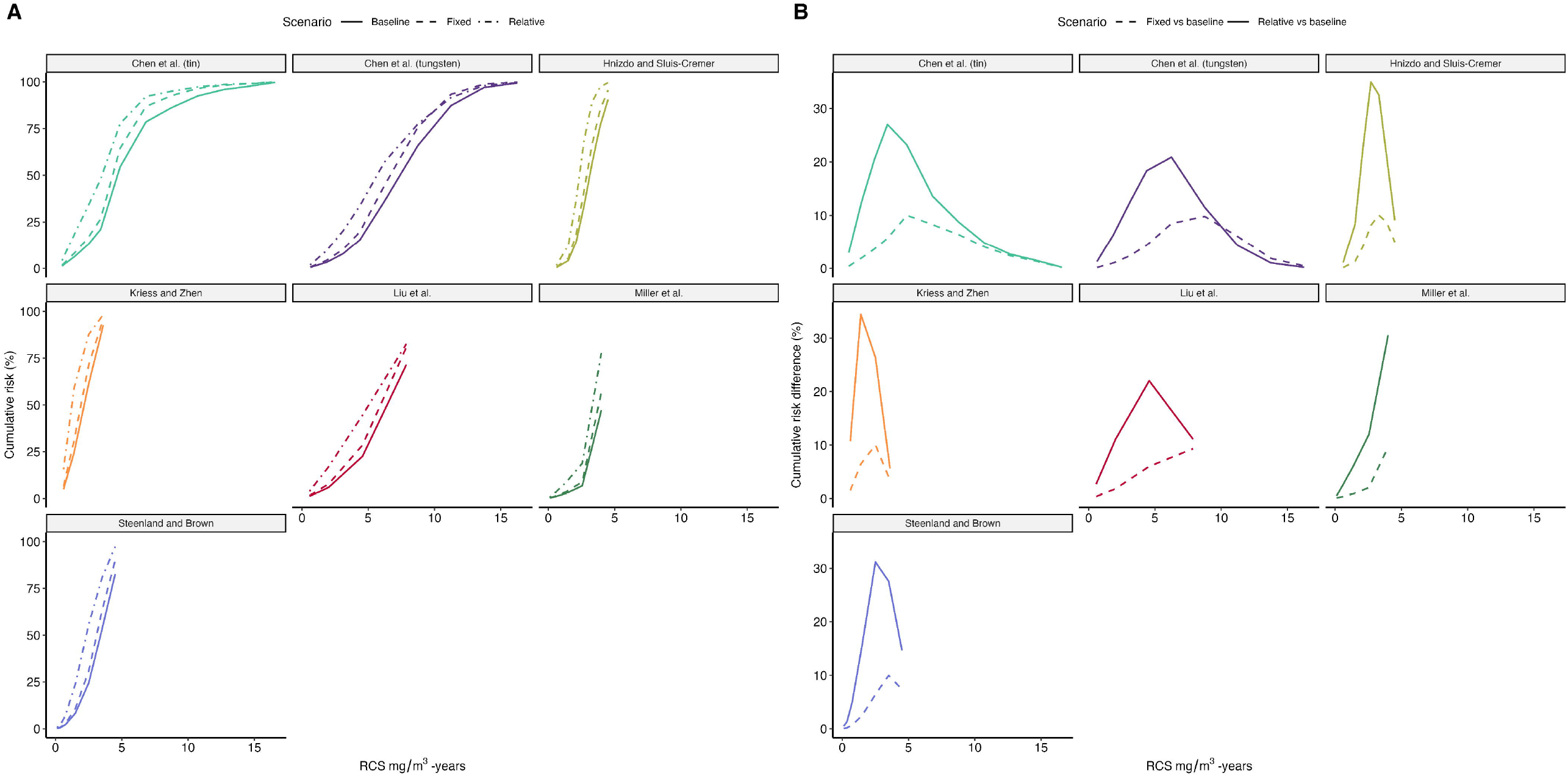
Comparison of cumulative risk of silicosis among studies of miners providing categorical data (n=6) as reported by the study (unadjusted; solid line) and modelled under an assumption of a fixed sensitivity of CXR compared to HRCT (fixed; dashed line) and a relative sensitivity of CXR compared to HRCT (relative; dot-dash line). Cumulative risks were calculated using the Steenland method. The studies are represented by the same positioning and colour in plots A and B. A. Comparison of cumulative risks according to cumulative silica exposure for eight cohorts across six studies. The solid line represents the unadjusted risk, dashed the fixed risk and dot-dash the relative risk. B. Re-representation of the data as the difference between unadjusted risk and adjusted scenarios of fixed and relative sensitivity, according to cumulative silica exposure. The solid line represents the difference between the fixed vs unadjusted risk, the dashed line represents the difference between the relative and unadjusted risk.

Among previously published mining cohorts, the cumulative risk of silicosis was higher in the fixed and relative sensitivity scenarios compared to the unadjusted estimate (Figure 1). This was most pronounced in the relative sensitivity scenario and in the lower range of exposures of approximately 6 mg/m^3^-years and under. As the cumulative risks of the studies approached 100% the differences reduced. The median cumulative risk at 4 mg/m^3^-years for the unadjusted, fixed and relative scenarios was 42% (IQR 24%, 62%), 51% (IQR: 20%, 71%) and 69% (IQR 45%, 87%), respectively (Supplementary table 2).

At lower cumulative RCS ranges (approximately <5 mg/m^3^-years) the dose-response curves for the three scenarios were similar (Supplementary figure 2 and 3). At higher ranges (>5 mg/m^3^-years), the profiles diverged; compared to the unadjusted estimated, the fixed scenario showed a relative increase, while the relative scenario decreased. In our secondary analysis with empirical knot choices a similar pattern was observed at the lower exposure ranges (Supplementary figure 4). At higher ranges, the difference between the fixed and unadjusted estimate narrowed, primarily as the unadjusted estimate was higher. The relative scenario still maintained an initial slight reduction in risk between 5 to 10 mg/m^3^-years, but then increased again. In all analyses, confidence intervals were wide above 5 mg/m^3^- years.

When the absolute risk of silicosis at 4 mg/m^3^-years was fixed at the LOESS-derived median cumulative risk estimates of respective scenarios, the absolute risk reduction in silicosis cases, when cumulative exposure was reduced to 2 mg/m^3^-years, in unadjusted, fixed and relative scenarios was 323 (95% CI: 298, 344) per 1000 miners, 409 (95% CI: 374, 434) per 1000 miners and 557 (95% CI: 451, 620) per 1000 miners, respectively (further exposure level estimates in Supplementary table 3). Secondary analysis results were similar (Supplementary table 4).

## Discussion

The poor sensitivity of CXR for silicosis has been a major limitation in our understanding of the relationship between cumulative silica exposure and silicosis risk [5]. We demonstrate that – when compared to the unadjusted risk – assuming either a fixed sensitivity for CXR or a sensitivity relative to the proportion of severe silicosis in the population results in substantial increases to the cumulative risk of silicosis among miners. This is most pronounced at lower ranges of cumulative exposure (e.g. under 6 mg-m^3^-years) which may be encountered more frequently in the workplace. Using previously published methods[1], when cumulative silica exposure is reduced from 4 to 2 mg/m^3^-years, substantially more cases were missed under the fixed and relative scenarios, compared to the unadjusted scenario.

A striking difference between the dose-response curves is the greater attenuation of risk at higher cumulative RCS exposures in the relative sensitivity scenario, compared to other scenarios. Possible reasons for attenuation at higher doses are well summarised in the article by Stayner et al [10]. Specifically, two factors may have influenced our observations. First, both the relative and fixed scenarios reached saturation of cases at 6 mg/m^3^-years (see Supplementary table 4), meaning all individuals in the population would (theoretically) have silicosis. Second, in the relative scenario, the sensitivity of CXR reduces with lower exposures. Thus, compared to the other scenarios, there is a marked increase in the *absolute risk* of silicosis in lower exposure groups, resulting in lower *relative* risks in higher exposure groups.

That both plausible, yet counterfactual, assumptions of fixed and relative sensitivity both demonstrate substantial differences from the unadjusted estimates lends weight to the conclusion that a true, clinically important underestimation of silicosis risk is likely. Given that the cumulative risk estimates of the individual studies we re-analysed [1] are often used when recommending or legislating exposure limits [2], estimating the impact of reduced CXR sensitivity is important.

In addition to underlying study limitations, our methods have limitations. We have used an HRCT reference standard for sensitivity; this may further underestimate silicosis risk compared to an autopsy reference. For our modelling, we used coefficients from models of 50-75 years old miners with over 20 years latency and exposure data with less than 2 mg/m^3^ intensity [7]. Although the Scottish coal miners study is comprehensive and well-regarded [2], our analysis may not be generalisable, particularly to populations with high intensity exposures [11].

Furthermore, the assumption that predicted probabilities from a logistic regression equate to population level estimates is simplistic. However, to the best of our knowledge, it is the only available data that provides estimates of ILO category risk according to cumulative exposure. Secondary analysis with different knot locations demonstrated that higher meta-analysis values were sensitive to knot placement, although subsequent ARRs were similar. These limitations demonstrate the reliance of our methods on our assumptions and highlight the multiple possible sources of error in our models. Thus, while our findings can be useful, we caution that the results should be viewed as illustrative rather than precise or predictive estimates.

In conclusion, currently reported estimates of silicosis risk according to cumulative silica exposure are likely significant underestimates due to the low sensitivity of CXR for silicosis.

## Data Availability

All data are available online at https://github.com/pjhowlett/da_silic_cxr and https://github.com/pjhowlett/silica_drma

https://github.com/pjhowlett/silica_drma

## Bibliography

1. Howlett P, Gan J, Lesosky M, Feary J. Relationship between cumulative silica exposure and silicosis: a systematic review and dose-response meta-analysis. Thorax. 2024 Sep 18;79(10):934–42.

2. OSHA. Occupational Exposure to Respirable Crystalline Silica. Final rule. [Internet]. Federal Register (USA); 2016 p. 16285–890. Report No.: 81(58). Available from: https://www.federalregister.gov/documents/2016/03/25/2016-04800/occupational-exposure-to-respirable-crystalline-silica

3. Mundt KA, Thompson WJ, Dhawan G, Checkoway H, Boffetta P. Systematic review of the epidemiological evidence of associations between quantified occupational exposure to respirable crystalline silica and the risk of silicosis and lung cancer. Front Public Health [Internet]. 2025 Feb 28 [cited 2025 Apr 1];13. Available from: https://www.frontiersin.org/journals/public-health/articles/10.3389/fpubh.2025.1554006/full

4. Ehrlich R, Murray J, Rees D. Subradiological silicosis. Am J Ind Med [Internet]. 2018 Nov [cited 2022 Oct 25];61(11):877–85. Available from: https://onlinelibrary.wiley.com/doi/10.1002/ajim.22909

5. Howlett P, Durairaj A, Lesosky M, Feary J. The diagnostic accuracy of chest Xray screening for silicosis: A systematic review, meta-analysis and modelling study [Internet]. medRxiv; 2025 [cited 2025 May 29]. p. 2025.05.28.25328086. Available from: https://www.medrxiv.org/content/10.1101/2025.05.28.25328086v1

6. Hnizdo E, Murray J, Sluis-Cremer GK, Thomas RG. Correlation between radiological and pathological diagnosis of silicosis: An autopsy population based study. Am J Ind Med [Internet]. 1993 Oct [cited 2022 Oct 25];24(4):427–45. Available from: https://onlinelibrary.wiley.com/doi/10.1002/ajim.4700240408

7. Miller BG, Hagen S, Love RG, Soutar CA, Cowie HA, Kidd MW, et al. Risks of silicosis in coalworkers exposed to unusual concentrations of respirable quartz. Occupational and Environmental Medicine [Internet]. 1998 Jan 1 [cited 2022 Oct 25];55(1):52–8. Available from: https://oem.bmj.com/lookup/doi/10.1136/oem.55.1.52

8. Buchanan D, Miller B, Soutar C. Research Report: Quantitative relationships between exposure to respirable quartz and risk of silicosis at one Scottish colliery. Edinburgh: Institute of Occupational Medicine; 2001.

9. Steenland K, Brown D. Silicosis among gold miners: exposure--response analyses and risk assessment. Am J Public Health [Internet]. 1995 Oct [cited 2022 Oct 25];85(10):1372–7. Available from: https://ajph.aphapublications.org/doi/full/10.2105/AJPH.85.10.1372

10. Stayner L, Steenland K, Dosemeci M, Hertz-Picciotto I. Attenuation of exposure-response curves in occupational cohort studies at high exposure levels. Scand J Work Environ Health [Internet]. 2003 Aug [cited 2023 Apr 13];29(4):317–24. Available from: http://www.sjweh.fi/show_abstract.php?abstract_id=737

11. Buchanan D, Miller BG, Soutar CA. Quantitative relations between exposure to respirable quartz and risk of silicosis. Occup Environ Med. 2003 Mar;60(3):159– 64.

