## Supplement for "Adjusting for the reduced sensitivity of CXR in the dose-response relationship between cumulative silica exposure and silicosis in miners"

**Author list and affiliations:**

1. *Patrick Howlett MBChB BSc MSc^1^ –
2. *Ashwin Durairaj BSc^1^ –
3. Jeffery Gan BSc^1^ -
4. Maia Lesosky BSc MSc PhD^1^ –
5. Johanna Feary BM BS MSc FRCP PhD^1,2^ –

Supplementary table 1. Example table of calculation of sensitivity, according to silica exposure, using the Scottish Coal miners logistic regression coefficients and our meta-regression formula. Silica exposures were converted from g/hr/m^3^ to mg/m^3^-years with the standard assumption that 1 year is 8 hours work a day for 270 days.

| Cumulative silica exposure (mg/m3-years) | Cumulative silica exposure (g/hr/m3 ) | Probability of ILO >= 1 disease (based on IOM formula) | Probability of ILO >= 2 disease (based on IOM formula) | Ratio of probabilities of >= ILO2 />= ILO 1 | Sensitivity (based on meta-regression formula) |
| --- | --- | --- | --- | --- | --- |
| 1 | 2.16 | 0.16 (0.09, 0.27) | 0.02 (0.01, 0.07) | 0.13 (0.06, 0.27) | 0.31 (0.23, 0.47) |
| 2 | 4.32 | 0.23 (0.12, 0.4) | 0.04 (0.01, 0.15) | 0.16 (0.07, 0.37) | 0.34 (0.24, 0.59) |
| 4 | 8.64 | 0.41 (0.19, 0.68) | 0.1 (0.01, 0.47) | 0.24 (0.07, 0.69) | 0.44 (0.25, 0.95) |
| 6 | 12.96 | 0.63 (0.29, 0.88) | 0.25 (0.02, 0.82) | 0.4 (0.09, 0.93) | 0.62 (0.26, 1) |
| 8 | 17.28 | 0.8 (0.42, 0.96) | 0.5 (0.04, 0.96) | 0.63 (0.11, 1) | 0.88 (0.28, 1) |
| 10 | 21.6 | 0.91 (0.55, 0.99) | 0.75 (0.08, 0.99) | 0.83 (0.14, 1) | 1 (0.32, 1) |

Supplementary table 2. Table of estimated cumulative risk of silicosis, calculated using the Steenland method, at corresponding cumulative silica exposures. Rows are separated according to unadjusted, fixed sensitivity and adjusted sensitivity scenarios. Summary rows of median and IQR are presented below each section. Presented studies are only mining cohorts.

|  | **Cumulative risk (%) at corresponding cumulative silica dose** | | | |
| --- | --- | --- | --- | --- |
|  | **1 mg/m^3^-years** | **2 mg/m^3^-years** | **4 mg/m^3^-years** | **10 mg/m^3^-years** |
| Unadjusted scenario | | | | |
| Chen et al. (tin) | 2.5 | 12.4 | 36.7 | 91.4 |
| Chen et al. (tungsten) | 1.1 | 3 | 13.1 | 77.3 |
| Hnizdo and Sluis-Cremer | 0 | 12.7 | 78.8 |  |
| Kriess and Zhen | 14.9 | 47 |  |  |
| Liu et al. | 2.4 | 6.0 | 19.5 |  |
| Miller et al. | 2.1 | 5.0 | 47.3 |  |
| Steenland and Brown | 3.8 | 15.2 | 66.7 |  |
| Median (IQR) | 2.4 (1.6, 3.1) | 12.4 (5.5, 13.9) | 42 (23.8, 61.9) | 84.3 (80.8, 87.9) |
| Fixed sensitivity scenario | | | | |
| Chen et al. (tin) | 3.6 | 16.1 | 44.1 | 96.4 |
| Chen et al. (tungsten) | 1.4 | 4.1 | 16.8 | 85.4 |
| Hnizdo and Sluis-Cremer | 0.2 | 16.6 | 87.1 |  |
| Kriess and Zhen | 18.8 | 55.9 |  |  |
| Liu et al. | 3.2 | 7.8 | 24.8 |  |
| Miller et al. | 2.7 | 6.5 | 56.9 |  |
| Steenland and Brown | 5.0 | 19.5 | 75.6 |  |
| Median (IQR) | 3.2 (2, 4.3) | 16.1 (7.2, 18) | 50.5 (29.6, 70.9) | 90.9 (88.2, 93.7) |
| Relative sensitivity scenario | | | | |
| Chen et al. (tin) | 11.5 | 29.5 | 61.1 | 97.4 |
| Chen et al. (tungsten) | 3.9 | 10.9 | 29.6 | 85.5 |
| Hnizdo and Sluis-Cremer | 3.4 | 34.3 | 98.9 |  |
| Kriess and Zhen | 37.4 | 78.7 |  |  |
| Liu et al. | 7.3 | 17 | 39.7 |  |
| Miller et al. | 6.5 | 14.2 | 77.8 |  |
| Steenland and Brown | 11.8 | 39.4 | 89.8 |  |
| Median (IQR) | 7.3 (5.2, 11.7) | 29.5 (15.6, 36.8) | 69.4 (45.1, 86.8) | 91.4 (88.5, 94.4) |

Supplementary table 3. Primary analysis. Comparison of the relative risk (as calculated in the dose-response meta-analysis) estimated number of silicosis cases per 1000 miners and absolute risk difference between levels of cumulative silica exposure

| Cumulative silica dose (mg/m^3^-years) | Risk ratio (95% CI) | Estimated number of silicosis per 1000 miners (95% CI) | Absolute risk difference compared to cumulative risk at 4 mg/m^3^-years per 1000 persons (95% CI) |
| --- | --- | --- | --- |
| Unadjusted scenario | | | |
| 1 | 0.07 (0.05, 0.1) | 29 (21, 42) | 391 (378, 399) |
| 2 | 0.23 (0.18, 0.29) | 97 (76, 122) | 323 (298, 344) |
| 4 | 1 | 420 | 0 (0, 0) |
| 6 | 2.25 (1.93, 2.62) | 945 (810, 1000) | -525 (-580, -391) |
| Fixed sensitivity scenario | | | |
| 1 | 0.05 (0.03, 0.08) | 25 (15, 40) | 480 (465, 490) |
| 2 | 0.19 (0.14, 0.26) | 96 (71, 131) | 409 (374, 434) |
| 4 | 1 | 505 | 0 (0, 0) |
| 6 | 2.77 (2.28, 3.37) | 1000 (1000, 1000) | -495 (-495, -495) |
| Relative sensitivity scenario | | | |
| 1 | 0.05 (0.02, 0.1) | 35 (14, 70) | 669 (634, 690) |
| 2 | 0.21 (0.12, 0.36) | 148 (85, 254) | 557 (451, 620) |
| 4 | 1 (1, 1) | 704 (704, 704) | 0 (0, 0) |
| 6 | 1.56 (1.26, 1.92) | 1000 (888, 1000) | -296 (-296, -183) |

Supplementary table 4. Secondary analysis with empirical knots. Comparison of the relative risk (as calculated in the dose-response meta-analysis) estimated number of silicosis cases per 1000 miners and absolute risk difference between levels of cumulative silica exposure

| Cumulative silica dose (mg/m^3^-years) | Risk ratio (95% CI) | Estimated number of silicosis per 1000 miners (95% CI) | Absolute risk difference compared to cumulative risk at 4 mg/m^3^-years per 1000 persons (95% CI) |
| --- | --- | --- | --- |
| Unadjusted scenario | | | |
| 1 | 0.07 (0.05, 0.1) | 29 (21, 42) | 391 (378, 399) |
| 2 | 0.21 (0.16, 0.26) | 88 (67, 109) | 332 (311, 353) |
| 4 | 1 | 420 | 0 |
| 6 | 1.56 (1.06, 2.31) | 655 (445, 970) | -235 (-550, -25) |
| Fixed sensitivity scenario | | | |
| 1 | 0.05 (0.03, 0.08) | 25 (15, 40) | 480 (465, 490) |
| 2 | 0.17 (0.13, 0.23) | 86 (66, 116) | 419 (389, 440) |
| 4 | 1 | 505 | 0 |
| 6 | 2.15 (1.15, 4) | 1000 (581, 1000) | -495 (-495, -76) |
| Relative sensitivity scenario | | | |
| 1 | 0.04 (0.02, 0.1) | 28 (14, 69) | 667 (625, 681) |
| 2 | 0.17 (0.1, 0.28) | 118 (69, 194) | 576 (500, 625) |
| 4 | 1 | 694 | 0 |
| 6 | 1.27 (0.5, 3.21) | 882 (347, 1000) | -188 (-306, 347) |

Supplementary figure 1. Modelled exposure-response relationship between cumulative silica exposure (mg/m³-years) and the sensitivity of CXR for detecting silicosis. The curve was derived by combining the mixed-effects linear meta-regression model with a logistic regression model from the Institute of Occupational Medicine (IOM) CXR sensitivity increases from a value of 0.29 at an exposure of 0 mg/m³-years to an with increasing silica exposure, reaching 1.0 (100% sensitivity) at 10.14 mg/m³-years. Sensitivity is equal to 0.29 when exposure is zero, because among the few cases estimated among the miners, 11% were of ≥ 2/1 category.

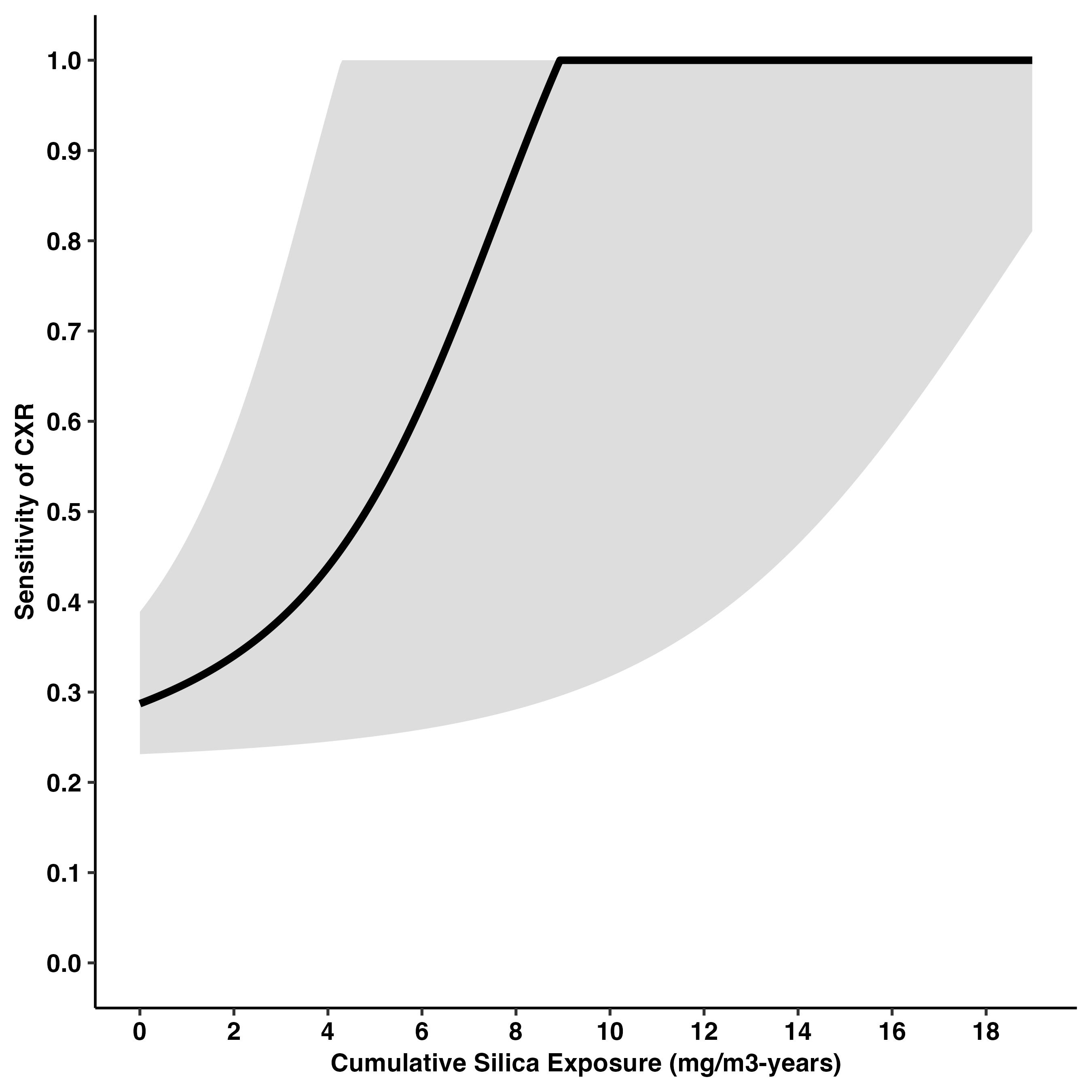

Supplementary figure 2. Log relative risk of silicosis according to cumulative silica exposure among mining cohorts. The colour of the point represents the study. The size of the point is relative to the standard error of the estimate. Unadjusted doses are not standardised (doses range 0.1 to 0.9 mg/m^3^-years). Plot A represents the unadjusted scenario (as in Howlett et al, 2024). Plot B represents the fixed sensitivity scenario. Plot C represents the relative sensitivity scenario.

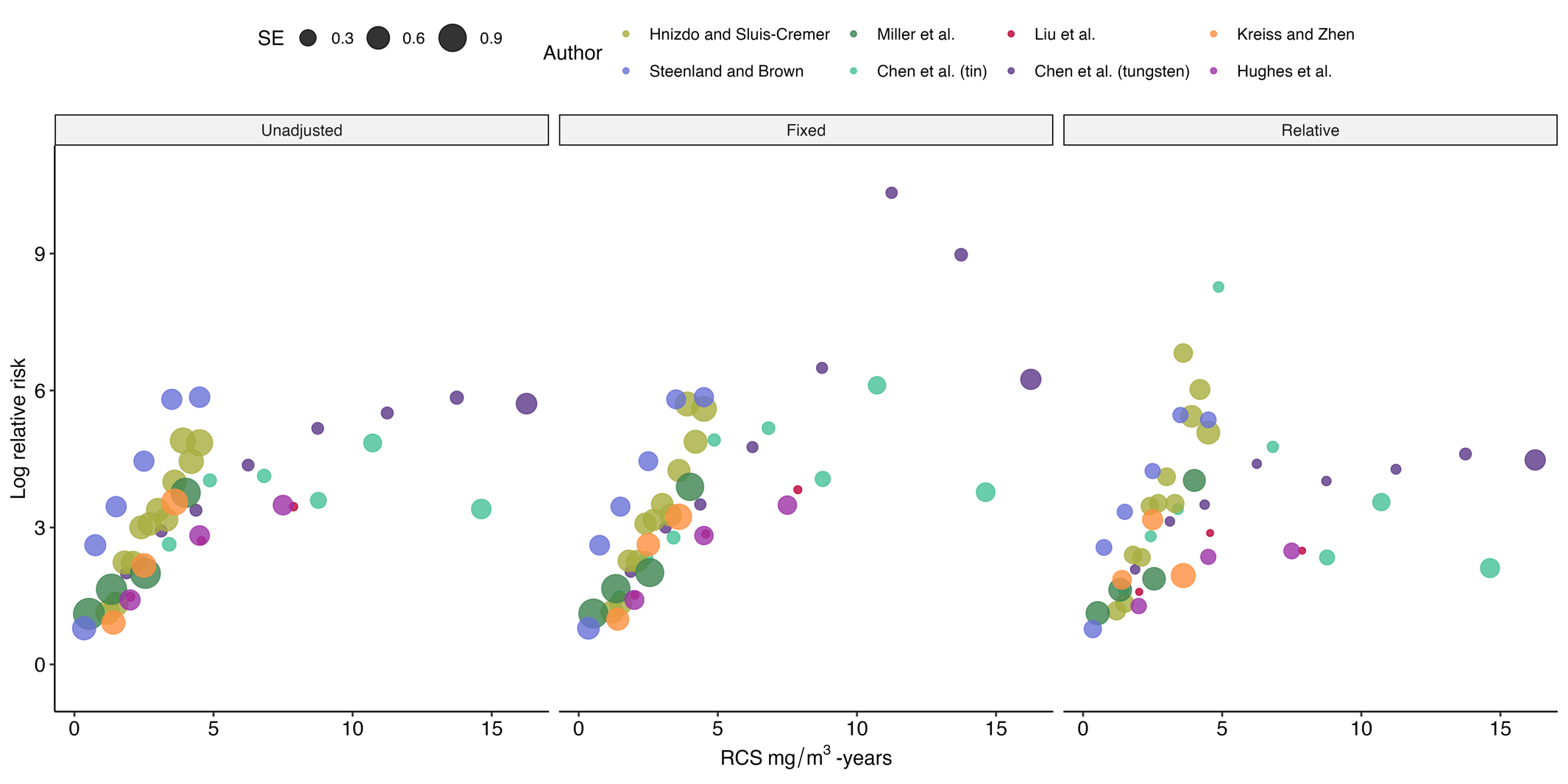

Supplementary figure 3. Dose-response meta-analysis of the relative risk of silicosis according to cumulative respirable crystalline silica (RCS) exposure using a restricted cubic spline model. The reference category with a relative risk of 1 is fixed at 4 mg/m^3^ -years, equivalent to 40 years working at 0.10 mg/m^3^. As the dose-response method would not allow for all persons in a category to achieve the outcome, 4 of the total 77 categories (across 8 cohorts) were combined. Four knots at default quantiles were chosen (0.5, 0.35, 0.65 and 0.95), equivalent to 0.4, 2.0, 3.9 and 12.0 mg/m^3^-years. Plot A represents the unadjusted scenario (as in Howlett et al, 2024). Plot B represents the fixed sensitivity scenario. Plot C represents the relative sensitivity scenario.

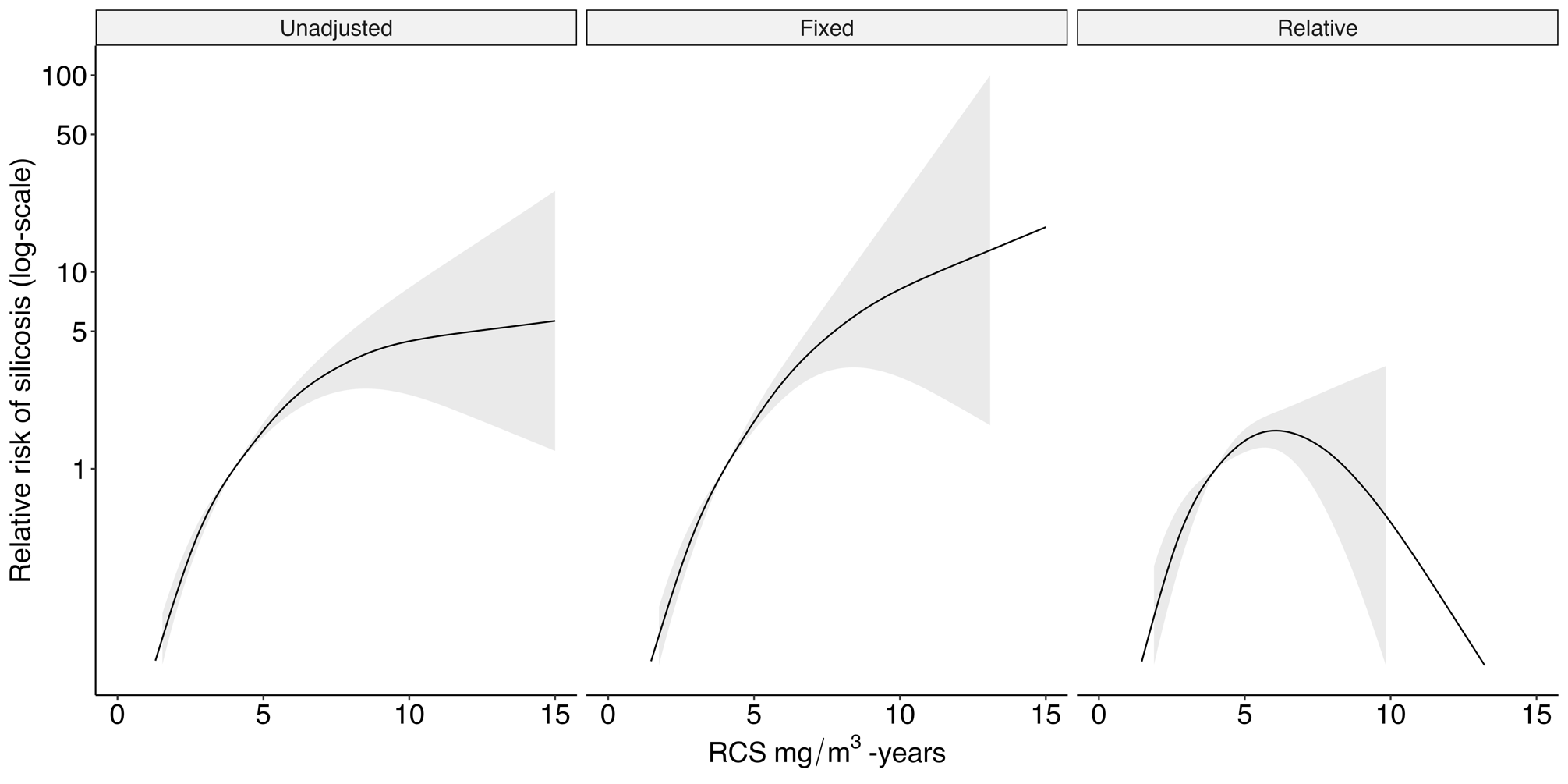

Supplementary figure 4. Secondary analysis with empirical knots. Dose-response meta-analysis of the relative risk of silicosis according to cumulative respirable crystalline silica (RCS) exposure using a restricted cubic spline model. The reference category with a relative risk of 1 is fixed at 4 mg/m^3^ -years, equivalent to 40 years working at 0.10 mg/m^3^. As the dose-response method would not allow for all persons in a category to achieve the outcome, 4 of the total 77 categories (across 8 cohorts) were combined. Four knots at empirical quantiles of 0.5, 3.5, 8 and 15.0 mg/m^3^-years were chosen. Plot A represents the unadjusted scenario (as in Howlett et al, 2024). Plot B represents the fixed sensitivity scenario. Plot C represents the relative sensitivity scenario.

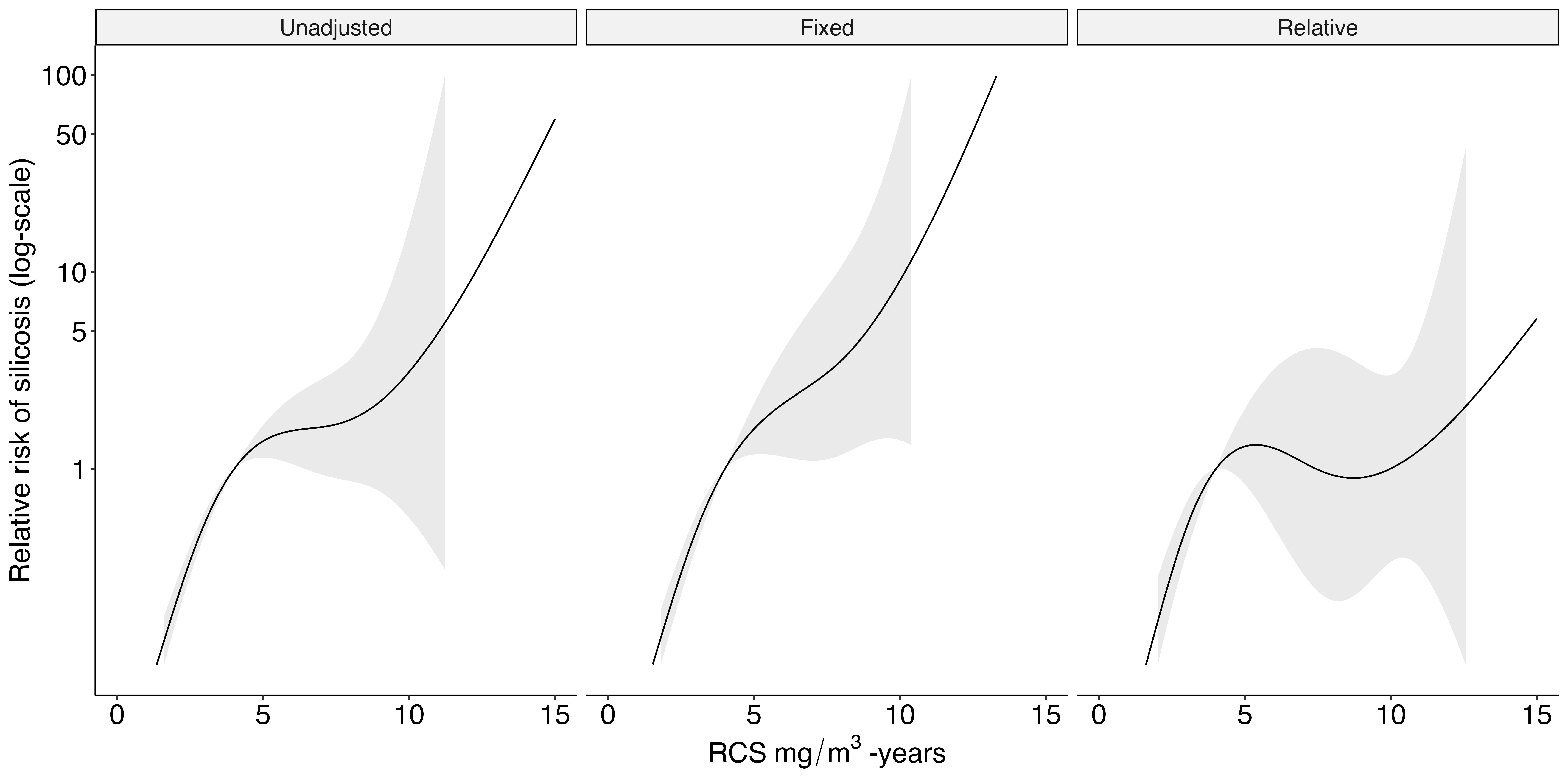
